# Creating a scalable CT yield metric for pulmonary embolisms in the emergency department using an open-source large language model

**DOI:** 10.64898/2026.01.13.26344087

**Authors:** Brittany C Wiseman, Henry Li, Edward Mason, Kevin Lonergan, Ross Mitchell, Jake Hayward

## Abstract

**Background:** CT scans are the gold-standard diagnostic test for pulmonary embolisms (PE). Despite stable PE prevalence, CT use is rising in emergency departments (EDs), suggesting test overuse. Current methods for measuring test yield are error-prone or not scalable, thus we tested the accuracy of an open-source, foundational large language model (LLM) for identifying PEs from free-text radiology reports.

**Methods:** Our retrospective diagnostic accuracy study used 10,173 CT-PE reports from 216 radiologists at 38 EDs across Alberta, Canada from April 2021-April 2023. Reports were classified as ‘PE present’, ‘PE absent’, or ‘Indeterminate’ by human labelers. An LLM (LLAMA-2-70B) was then prompt-engineered to label the reports. Label accuracy was compared against ICD-10-CA codes and a rule-based natural language processing (NLP) algorithm (ChartExtract; University of Toronto). Descriptive statistics were performed to analyze results.

**Results:** 1070 (11.8%) reports were PE positive. The LLM achieved an Area Under the Curve (AUC) of 99.1%, outperforming both ICD-10-CA (AUC=90.6%) and ChartExtract (AUC=86.5%), while demonstrating a 16-25% higher sensitivity (LLM: sensitivity=98.8%, specificity=99.1%; ICD-10-CA: sensitivity=82.0%, specificity=99.1%; ChartExtract: sensitivity=73.6%, specificity=99.4%). The LLM took an average of 143 milliseconds to label each report and produce a paragraph justifying the classification.

**Conclusions:** Open-source, foundational LLMs are an accurate and scalable method for interpreting radiology reports and identifying PEs from ED data. If IT resources are available, this is a cost-effective approach to quality metric derivation for diagnostic processes in large health systems.

## 1. Introduction

Timely diagnosis of pulmonary embolism (PE) is crucial for reducing mortality and morbidity[1] and a computed tomography scan for pulmonary embolism (CT-PE) is the diagnostic test of choice[2]. Despite their efficacy, CT scans are costly, time-consuming, and carry potential health risks, including radiation exposure, prolonged emergency department (ED) stays, and increased healthcare expenditure[3].

In recent decades, CT scan use has increased while the incidence of PE has remained stable, suggesting potential test overuse[4–7]. To monitor and improve test appropriateness, it is important that health systems track test yield, or the proportion of CT scans that are positive. This is challenging to measure using structured administrative data, including diagnostic codes, which are prone to inaccuracies[8–10]. Theoretically, the best approach might be to extract information directly from free text radiology reports, but this usually requires manual chart reviews that are too resource intensive for use in large populations[7,11].

Natural language processing (NLP) offers a potential method for extracting information from radiology reports[12,13]. While some NLP algorithms rely heavily on humans for derivation (i.e, rule-based methods), large language models (LLMs) have now emerged as highly flexible and efficient options for NLP that require minimal human input[13–15]. Many of these models, referred to as “foundational” LLMs, have been pre-trained on a vast corpora of text spanning a wide range of subjects and can be applied to multiple downstream applications without the need for model retraining[16]. A subset of foundational LLMs are available open-source and free of charge.

The objective of this study was to assess whether a foundational, open-source LLM (LLaMA-2, Meta Inc., Menlo Park, CA) could accurately extract information from CT-PE radiology reports. If successful, this approach could facilitate a range of clinical applications, including rapid, scalable, and cost-efficient test-yield metrics, forming a basis for scalable quality improvement in large and distributed populations[17].

## 2. Methods

### 2.1 Study Design

We conducted a retrospective test accuracy study to analyze 10,173 CT-PE radiology reports written by 216 radiologists from 38 different EDs across Alberta, Canada between April 2021 and April 2023.

### 2.2 Study Setting and Population

Alberta covers 661,848 km² and has a population of approximately 4.4 million[18]. Alberta Health Services (AHS) began implementing a province wide EMR (Epic Systems Inc. Verona SI) in 2019. We enrolled a convenience sample of adult ED patients who underwent a CT-PE scan at one of the 38 EDs where the provincial EMR was operational between April 1st, 2021, and April 1st, 2023. This study was approved by research ethics (Pro00127814). The randomly selected subject group was 53.3% female, with an average age of 61.

### 2.3 Large Language Model Infrastructure

One of the authors developed a custom LLM prompt-tuning software application using Python (v3.10). This software employs a browser-based user interface based on the Gradio library and was deployed on a Graphics Processing Unit (GPU) server (Hyperplane, Lambda Labs Inc.) equipped with 8 x A100 GPUs (Nvidia Inc.). The system also includes 2 x EPYC 7763 64-core Central Processing Units (CPUs) (AMD Inc.), 3TB of DDR4-3200 Random Access Memory (RAM), and 6 x 15TB Non-Volatile Memory Express (NVMe) solid-state drives. Additionally, author RM developed a Python (v3.10) algorithm for parallel processing of medical texts using an LLM. We utilized an open-source LLM based on LLAMA-2-70B (upstage/Llama-2-70b-instruct available at https://huggingface.co/).

### 2.4 Data Sources

Data was obtained from the AHS Enterprise Data Warehouse (EDW), a central repository for EMR and other administrative data in Alberta, which includes the EPIC Clarity database supporting the AHS EMR (i.e. Connect Care). All Alberta residents are provided a unique identifier under the Alberta Health Care Insurance Plan (AHCIP) allowing for deterministic linkage of different administrative data sources and robust databases for this study. Personal health information (PHI) within the radiology reports was anonymized in compliance with AHS guidelines.

### 2.5 Labeling Pipeline

Study team members categorized each CT report in the initial set of radiology reports (N=10,173) as ‘PE present,’ ‘PE absent,’ or ‘Indeterminate’. The ‘indeterminate’ category refers to CT reports in which the radiologist made explicit mention of insufficient images for an accurate diagnosis (ex. motion artifact) and does not refer to reports that were incomplete or had missing text (which were later removed). A random sample of 200 reports was used to train team members on the labeling task and to refine the labeling pipeline. After training, interrater reliability for two independent human labelers was near perfect (κ=0.973, n=200). The remaining reports were then labeled by a single study team member.

### 2.6 Prompt Engineering

Figure 1 visually depicts the dataset splits used for prompt engineering. This consisted of a two-phase approach with iterative testing and adjustment of an initial prompt candidate in sequentially larger sub-samples of the data. In Phase 1, adjustments to the prompt were based on discrepancies between the LLM and human labels, using a random sample of 100 reports. In Phase 2, the refined prompt was tested using another set of 1000 reports.

**Figure 1.**
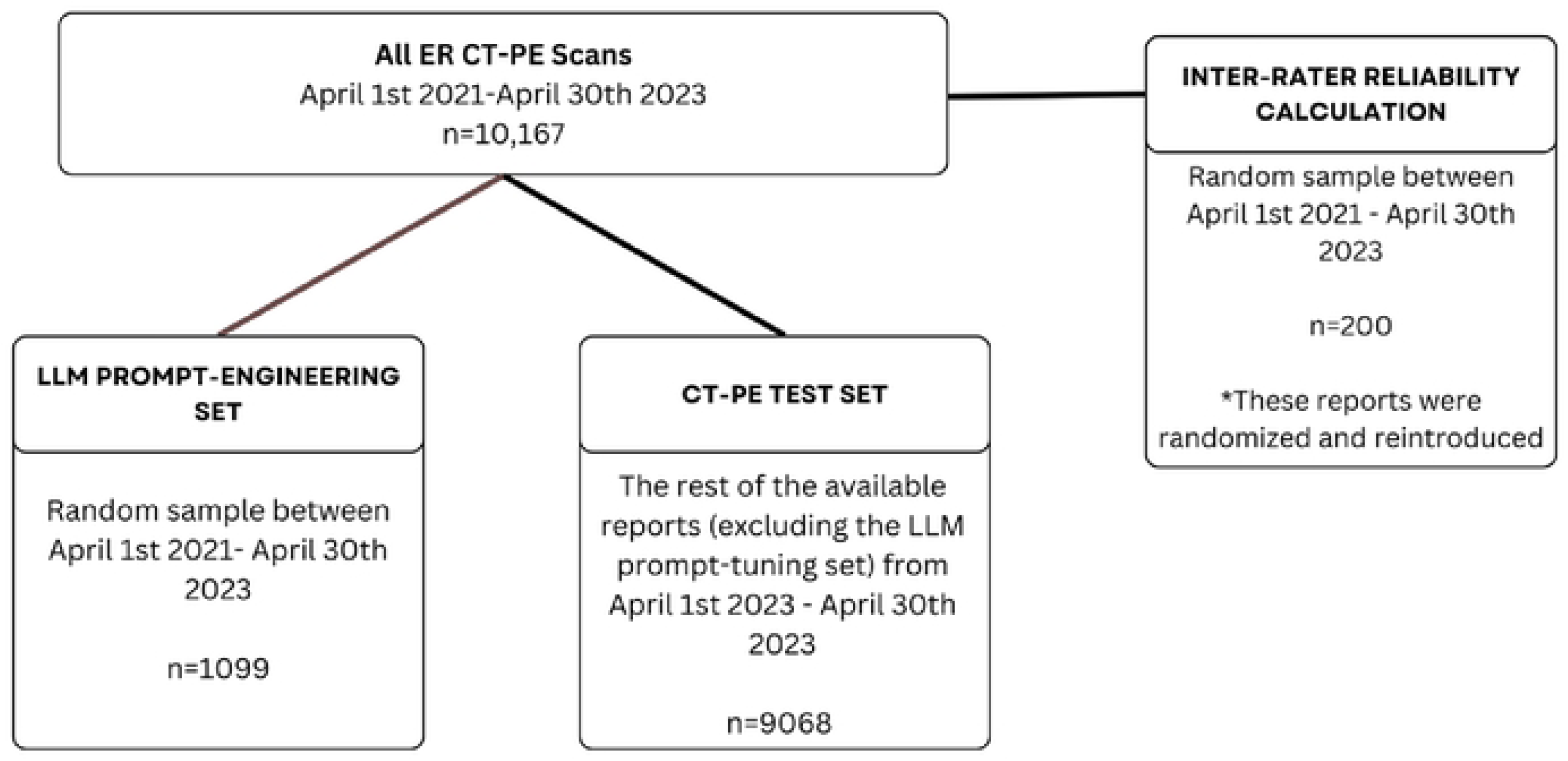
Cohort derivation

Phase 1: The initial candidate prompt (see supplementary materials) was tested and refined using a random sample of 100 reports. For each prompt version, a team member reviewed cases where the LLM and human labels disagreed, and the information gained was then used to adjust the prompt in the next iteration. This process was carried out with the same 100 reports until negligible improvements were observed with further changes to the prompt.

Phase 2: The refined prompt from Phase 1 underwent the same process of iterative testing and revision using a larger randomly selected sample of 1000 reports. Subsequently, 1 report was removed from the 1000 as it was determined to have incomplete/missing information that precluded categorization, yielding 999 total reports for prompt testing. As in Phase 1, when no further improvements in accuracy were achieved through prompt revisions, the process was stopped.

The final prompt is provided below (see Supplementary Materials for the full sequence of prompt development), which included instructions to generate an explanation/rationale for each label; this helped the authors interpret discrepancies between LLM and human labels. The final prompt consisted of two parts: the system prompt (high level instructions) and the user prompt (detailed instructions):

> System prompt: “You are an experienced nurse skilled at extracting information from clinical notes. Use the following clinical note to answer the user’s questions. Be brief. If the answer is not provided in the following clinical note, simply reply that the answer to the question is unknown. Do not try to make up an answer.”

> User prompt: “Do the patient’s lungs have a pulmonary embolism (PE), thromboembolic disease, thrombus, clot burden, a filling defect, or sequelae of pulmonary emboli? The correct answer is ‘yes’ if any of these conditions are chronic or present in a small, trace, or low extent. The correct answer is ‘no’ if the exam is normal or if any of these conditions occur outside the lungs. If none of these findings are present, then the correct answer is not ‘unknown’ it is ‘no’. Answer with a single word, one of ‘yes’ ‘no’ or ‘unknown’. Provide this answer on a line by itself. Then explain your reasoning.”

### 2.7 Error Analysis

The 1,100 reports used for prompt-engineering were removed, leaving 9,073 unseen reports used for LLM testing. Two team members manually reviewed discrepant human and LLM labels to discern errors attributable to humans versus the LLM. Disagreements were resolved through consensus via discussion). Subsequently, 5 reports were removed from the test set due to incomplete/missing information and insufficient data for labeling, for example, typos, missing words, or incomplete sentences. The final test set therefore included 9,068 reports. Human labeling errors (identified through analysis of LLM disagreements) were corrected prior to final analysis (Figure 2).

**Figure 2.**
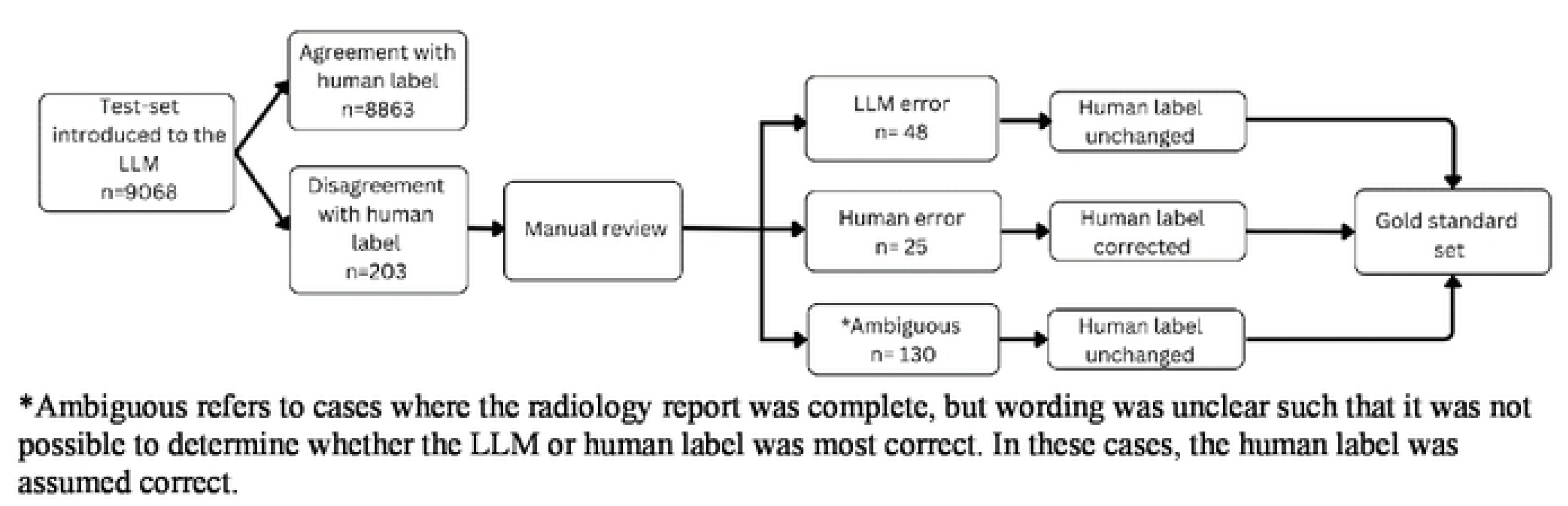
Error analysis and correction

### 2.8 Comparison to Other Methods

The LLM’s performance was first tested against ICD-10-CA diagnostic codes derived from the corresponding ED visit (I26.9: pulmonary embolism without cor pulmonale, and I26.0: pulmonary embolism with cor pulmonale)[8–10]. We then compared performance to an open-source traditional NLP algorithm developed at the University of Toronto (ChartExtract[19]), which uses a rule-based approach and was selected for its relevance to the Canadian context. To further investigate the relationship between diagnostic codes and radiology reports, we described the distribution of ICD codes for the subgroup of ED visits where the CT scan was positive for PE but no ICD-10-CA code for PE was recorded.

### 2.9 Data Analysis

Patient demographics and visit characteristics were summarized using descriptive statistics with means and proportions for continuous and categorical variables, respectively. We calculated area under the receiver operator curve (AUC), sensitivity, specificity, positive predictive value (PPV) and negative predictive value (NPV) for each labeling method (LLM vs. ICD-10-CA vs. ChartExtract). For the primary analysis, indeterminate labels were grouped with the ‘PE absent’ category, consistent with previous research[20]. Sensitivity analyses tested different approaches to the analysis of indeterminate labels, for both human and LLM, including omitting them altogether (Supplemental materials). The sample size incorporated all available radiology reports for the time period, ensuring adequate power and significance.

## 3. Results

### 3.1 Population characteristics

Patient characteristics for all datasets are summarized in Table 1. Prompt engineering (n=1099) and test (n=9068) sets were well-balanced across all patient variables. The overall sample comprised 10,173 CT scans performed mostly in females (53.3%). The average age was 61 years old. Overall, 1070 (11.8%) CT-PE scans were positive for PE and 65 (0.7%) were indeterminate. Most visits were for ambulatory patients (56.4%), predominantly at rural hospitals (39.9%), with moderate to high triage acuity scores (Triage 2=48.1% and Triage 3=40.0%;1=highest acuity, 5=lowest acuity). The most common presenting complaints were shortness of breath (31.7%) and cardiac chest pain (21.4%).

**Table 1.** Cohort characteristics.

| **Characteristic** | **All CT PE Scans** | **LLM Prompt-Engineering Set** | **CT-PE Test Set** |
| --- | --- | --- | --- |
| **Number of scans** | 10,167 | 1099 | 9068 |
| **Visit Year N (%)** | 2021: 2333 (22.9%) 2022: 4649 (45.7%) 2023: 3185 (31.3%) | 2021: 253 (23.0%) 2022: 524 (47.7%) 2023: 322 (29.3%) | 2021: 2080 (22.9%) 2022: 4125 (45.5%) 2023: 2863 (31.6%) |
| **Average age: mean (SD)** | 60.6 (20.5) | 60.1 (18.2) | 60.7 (20.8) |
| **Age Category N (%)** | <50: 2820 (27.7%) >50: 7347 (72.2%) | <50: 293 (26.7%) >50: 806 (73.3%) | <50: 2527 (27.9%) >50: 6541 (72.1%) |
| **Sex N (%)** | F: 5421 (53.3%) M: 4746 (46.7%) | F: 586 (53.3%) M: 513 (46.7%) | F: 4835 (53.3%) M: 4233 (46.7%) |
| **Hospital Type N (%)** | Urban: 3282 (32.3%) Regional: 778 (7.7%) Rural: 4060 (39.9%) Missing: 2047 (20.1%) | Urban: 362 (32.9%) Regional: 87 (7.9%) Rural: 426 (38.8%) Missing: 224 (20.4%) | Urban: 2920 (32.2%) Regional: 691 (7.6%) Rural: 3634 (40.1%) Missing: 1823 (20.1%) |
| **Arrival Mode N (%)** | EMS: 3908 (38.4%) Ambulatory: 5735 (56.4%) Missing: 524 (5.2%) | EMS: 436 (39.7%) Ambulatory: 610 (55.5%) Missing: 53 (4.8%) | EMS: 3472 (38.3%) Ambulatory: 5125 (56.5%) Missing: 471 (5.2%) |
| **\*Triage Level N (%)** | 1: 431 (4.2%)<br>2: 4895 (48.1%)<br>3: 4026 (40.0%)<br>4: 300 (3.0%)<br>5: 272 (2.7%)<br>Missing: 497 (4.9%) | 1: 58 (5.3%)<br>2: 514 (46.8%)<br>3: 441 (40.1%)<br>4: 33 (3.0%)<br>5: 5 (0.5%)<br>Missing: 48 (4.4%) | 1: 373 (4.1%)<br>2: 4381 (48.3%)<br>3: 3585 (39.5%)<br>4: 267 (2.9%)<br>5: 267 (2.9%)<br>Missing: 449 (5.0%) |
| **\*\*Presenting Complaint N (%)** | SOB: 3119 (30.7%)<br>CCP: 2172 (21.4%) NCCP: 820 (8.1%)<br>GW: 351 (3.5%)<br>S: 413 (4.1%) | SOB: 318 (28.9%)<br>CCP: 226 (20.6%) NCCP: 93 (8.5%)<br>GW: 47 (4.3%)<br>S: 51 (4.6%) | SOB: 2801 (30.9%)<br>CCP: 1946 (21.5%) NCCP: 727 (8.0%)<br>GW: 304 (3.4%)<br>S: 362 (4.0%) |

\*Triage level 1= highest acuity, 5= lowest acuity (Canadian Triage and Acuity Scale) \*\*SOB: Shortness of breath, CCP: Cardiac chest pain, NCCP: Non-cardiac chest pain, GW: general weakness, S: syncope/ pre-syncope

### 3.2 Computation Time

The parallel batch processing software running on the GPU server required 21 minutes and 37 seconds to process 9,073 reports (average of 143ms per report), including the time to label each report and generate a short explanatory paragraph.

### 3.3 Human vs. LLM Error

Figure 2 includes a description of labeling error analysis. There were 203 disagreements between human and LLM labels (2.0%). Following manual review, 48 of these were attributed to LLM error, 25 to human error, and 130 of the disagreements were too ambiguous to categorize accurately. Assuming a negligible rate of dual-labeling error (i.e. both human and LLM are wrong) this equates to error rates of 0.3% and 0.5% for human and LLM, respectively.

### 3.4 Comparative Accuracy

Table 2 displays the comparative accuracy of candidate methods for identifying PEs. The LLM demonstrated the highest accuracy, achieving a sensitivity of 98.9%, specificity of 99.5%, and an AUC of 99.1%. This outperformed ICD-10-CA diagnostic codes and ChartExtract, which achieved sensitivities of 82.0% and 73.6%, respectively. All three methods achieved specificities above 99.0%. Performance did not change significantly in various sensitivity analyses (see Supplementary Table 2).

**Table 2.** Comparative accuracy of labeling techniques.

|  | **LLM** | **ChartExtract** | **ICD-10-CA** |
| --- | --- | --- | --- |
| **Sensitivity** | 0.988 | 0.736 | 0.820 |
| **Specificity** | 0.995 | 0.994 | 0.991 |
| **PPV** | 0.961 | 0.940 | 0.928 |
| **NPV** | 0.998 | 0.966 | 0.976 |
| **Positive LR** | 183.740 | 117.653 | 96.403 |
| **Negative LR** | 0.012 | 0.266 | 0.182 |
| **AUROC** | 0.991 | 0.865 | 0.906 |

PPV: positive predictive value, NPV: negative predictive value, LR: likelihood ratio, AUROC: Area Under the Receiver Operating Characteristics (ROC) curve.

### 3.5 Disagreement with Diagnostic Codes

Diagnostic codes in administrative data are known to lack sensitivity for identifying PEs[8]. Among ED visits with a positive CT-PE but no corresponding diagnostic code, the most common code was for hypertension (I100; 8.6%), followed by pneumonia (J189; 4.8%), asphyxia and hypoxemia (R090; 4.0%), dyspnea unspecified (R060; 3.8%), and unspecified chest pain (R074; 3.5%). Additional codes are detailed in Table 3.

**Table 3.** Top non-PE diagnostic codes used for instances of PE positive CT scans.

| **ICD-10-CA Code** | **ICD-10-CA code description** | **Count** | **Percent (%)** |
| --- | --- | --- | --- |
| **I100** | Hypertension | 54 | 8.60% |
| **J189** | Pneumonia, unspecified organism | 30 | 4.78% |
| **R090** | Asphyxia and hypoxemia | 25 | 3.98% |
| **R060** | Dyspnea, unspecified | 24 | 3.82% |
| **R074** | Chest pain, unspecified | 22 | 3.50% |
| **I500** | Congestive Heart Failure | 16 | 2.55% |
| **I4890** | Atrial fibrillation and atrial flutter, unspecified | 16 | 2.55% |
| **U071** | COVID-19: Confirmed by laboratory testing | 15 | 2.39% |
| **E119** | Type 2 diabetes mellitus without complications | 12 | 1.91% |
| **J440** | Chronic obstructive pulmonary disease with (acute) lower respiratory infection | 11 | 1.75% |

## 4. Discussion

We demonstrate that a prompt-engineered, open-source, foundational LLM (LLaMA-2-70B) outperforms traditional methods in identifying PEs from routinely collected administrative data. Compared to diagnostic codes and a rule-based NLP algorithm (ChartExtract), the LLM improves sensitivity by 16–25%, identifying an additional 192–300 PEs per 10,000 scans (PE prevalence ∼12%). Our approach is scalable across large health systems, processing 9,073 reports in under 22 minutes.

NLP-based radiology report processing is crucial, as diagnostic codes are often inaccurate, leading to misestimates of disease prevalence [8–10]. This issue worsens with data drift - shifts in the validity and accuracy of structured EMR data over time[21,22]. We show that rule-based NLP algorithms perform poorly, even worse than diagnostic codes, underscoring the potential of LLMs to extract clinically meaningful insights from health records. Beyond radiology reports, LLMs could unlock vast amounts of information from EMRs, enhancing the utility of routinely collected structured data[23,24].

More accurate disease identification could enable a new generation of quality metrics better suited to informing clinical care[25]. Tracking sentinel diagnoses such as PE, appendicitis, and stroke provides essential information to address quality gaps and drive improvements [25,26]. Research also benefits, as routinely collected administrative datasets can facilitate more representative study populations without manual chart review [27]. We demonstrate that even early-generation foundational LLMs, without fine-tuning for medical contexts, appear to achieve near-perfect performance in disease identification. This breakthrough paves the way for large-scale disease tracking across diverse populations, unlocking important opportunities for clinical and epidemiological research [28,29].

Our experience provides practical insights into the trade-offs, technical barriers, and implementation challenges health systems face when applying generative AI to real-world clinical problems. Our multidisciplinary team—comprising computer scientists, analysts, and clinicians—found that close collaboration was essential to align clinical objectives, technical requirements, and resource constraints, particularly for labeling and prompt engineering. The cost of applying LLMs in clinical settings is therefore substantial (although not directly measured in our study), potentially limiting the range of feasible applications. This highlights the importance of strategic planning and context-specific prioritization, tempering the notion of generative AI as a catch-all solution for EMR data extraction.

Data privacy and security are critical concerns, as many third-party LLM services (e.g., OpenAI) require patient data to be transferred to external servers—often temporarily but sometimes for proprietary model training. Organizations must navigate local laws, regulations, and data stewardship policies to ensure compliance and, where guidelines are lacking, actively contribute to policy development. To adhere to our organization’s data-sharing restrictions, we hosted our open-source LLM in a closed environment behind a secure firewall. While this prevented external data transfer—a key advantage—it also placed significant demands on local IT infrastructure. For small-scale implementations, in-house solutions may suffice, but scaling to larger models or broader disease applications could exceed local computational capacity[17]. As LLM adoption grows and models become more accessible and commoditized, healthcare organizations may increasingly rely on cloud and model service providers (e.g., Microsoft Azure, AWS). This shift introduces new privacy and security vulnerabilities, emphasizing the need for robust safeguards in data sharing and access control.

Our prompt-engineering approach was novel and offers insights for future studies. The process involved: 1) Iteratively testing and refining a candidate prompt on a small labeled subset (e.g., 200 reports in this study); 2) Expanding to progressively larger labeled subsets until achieving sufficient accuracy or exhausting human resources. Using this method, we developed a high-performing prompt while labeling just over 10% of our total sample (1,000 reports).

However, as mentioned above, prompt engineering required substantial effort in assembling a team of labeling experts, posing a persistent constraint on future applications. To scale effectively, reducing reliance on human labeling and leveraging automated intelligence labelling technologies will be essential [24,31].

An unexpected advantage of our approach was its ability to detect human labeling errors that might have otherwise gone unnoticed. While the LLM was not inherently more accurate than humans, it made different types of errors, suggesting a synergistic effect—achieving greater accuracy together than individually[27]. Our findings also call into question the future role of traditional diagnostic codes in quality metric derivation. ICD codes often capture symptoms (e.g., unspecified chest pain, shortness of breath) rather than definitive diagnoses. However, they remain a quick and inexpensive tool. Organizations must weigh the trade-offs between investing in labeling and LLM technologies versus relying on the convenience and lower cost of traditional coding.

## 5. Limitations

Our study has several limitations. The ChartExtract NLP tool requires re-tuning for each new dataset, as indicated by its authors [19], which we did not perform. Re-tuning would have required significant effort with little likelihood of surpassing LLM performance. Our calculated error rates may not be perfectly accurate, as we did not account for dual errors—cases where both the human and LLM were incorrect—since these are difficult to identify without manual chart review. However, given the low observed error rates, such events are likely rare and unlikely to meaningfully impact our results. The LLaMA-2 LLM’s performance may not generalize across all settings, particularly in different countries and languages, and our final prompt may also not be applicable to other LLMs [32]. However, our dataset was diverse, including hospitals across a large geographic area, with both rural and urban facilities. Notably, in previous head-to-head comparisons, pay-for-use LLMs (e.g., ChatGPT) have outperformed open-source models like LLaMA-2 on certain tasks [23]. Yet, given the near-perfect performance of our model in PE identification, the added cost of proprietary models may offer little return on investment.

## 6. Conclusions

In summary, our study demonstrates the feasibility of using open-source, foundational LLMs for disease identification in EMR unstructured data. This approach effectively estimates CT PE test yield in the ED and can be readily applied to other diseases and radiological tests. Even at this early stage of development for LLM technology, open-source, general models are sufficiently accurate for this specialized task, offering a scalable and cost-effective alternative to manual chart reviews or fine-tuning LLMs. By the time of publication, newer and more advanced LLMs will be available, further enhancing their utility in future work. With thoughtful implementation, these groundbreaking tools can support quality improvement and research initiatives, particularly for large-scale projects requiring extensive free-text processing.

## 7. Acknowledgments

The authors were supported through an Undergraduate Summer Studentship from the Alberta Health Services (AHS) Emergency Strategic Clinical Network (ESCN) and by research chair positions from Alberta Health Services (AHS) and the Canadian Institute for Advanced Research (CIFAR), as well as funding from the University Hospital Foundation (UHF), and Alberta Machine Intelligence Institute (Amii).

## Data Availability

Data cannot be shared publicly due to patient confidentiality. Unidentified files can be made available for researchers who meet criteria for access to confidential data through Alberta Health Services by contacting the corresponding author, Dr Jake Hayward.

## Supplemental Files

Supplemental Table 1. Confusion matrix for LLM vs manual review after human corrections

Supplemental Table 2. Sensitivity analysis of indeterminate labels

Supplemental Table 3. Distribution of visits across hospitals in Alberta

## References

1. Barco S, Valerio L, Ageno W, Cohen A, Goldhaber S, Hunt B, et al. Age-sex specific pulmonary embolism-related mortality in the USA and Canada, 2000–18: an analysis of the WHO Mortality Database and of the CDC Multiple Cause of Death database. Lancet Respir Med. 2021;9(1):33–42. doi:10.1016/S2213-2600(20)30417-3(Barco et al., 2021)

2. Moore AJE, Wachsmann J, Chamarthy MR, Panjikaran L, Tanabe Y, and Rajiah P. Imaging of acute pulmonary embolism: an update. Cardiovasc Diagn Ther. 2018;8(3):225–243. doi:10.21037/cdt.2017.12.01

3. Cao CF, Ma KL, Shan H, Liu T, Zhao S, Wan Y, et al. CT Scans and Cancer Risks: A Systematic Review and Dose-response Meta-analysis. BMC Cancer. 2022;22(1):1238. doi:10.1186/s12885-022-10310-2

4. Chung JH, Duszak R Jr, Hemingway J, Hughes D and Rosenkrantz A. Increasing Utilization of Chest Imaging in US Emergency Departments From 1994 to 2015. J Am Coll Radiol. 2019;16(5):674–682. doi:10.1016/j.jacr.2018.11.011

5. Feng LB, Pines JM, Yusuf HR, and Grosse S. U.S. Trends in Computed Tomography Use and Diagnoses in Emergency Department Visits by Patients With Symptoms Suggestive of Pulmonary Embolism, 2001–2009. Acad Emerg Med. 2013;20(10):1033–1040. doi:10.1111/acem.12221

6. Kline JA, Garrett JS, Sarmiento EJ, Strachan C, and Courtney DM. Over-Testing for Suspected Pulmonary Embolism in American Emergency Departments. Circ Cardiovasc Qual Outcomes. 2020;13(1):e005753. doi:10.1161/CIRCOUTCOMES.119.005753

7. Venkatesh AK, Agha L, Abaluck J, Rothenberg C, Kabrhel C, and Raja A. Trends and Variation in the Utilization and Diagnostic Yield of Chest Imaging for Medicare Patients With Suspected Pulmonary Embolism in the Emergency Department. Am J Roentgenol. 2018;210(3):572–577. doi:10.2214/AJR.17.18586

8. Burles K, Innes G, Senior K, Jang E and McRae A. Limitations of pulmonary embolism ICD-10 codes in emergency department administrative data: let the buyer beware. BMC Med Res Methodol. 2017;17(1):89. doi:10.1186/s12874-017-0361-1

9. Peng M, Eastwood C, Boxill A, Jolley RJ, Rutherford L, Carlson K et al. Coding reliability and agreement of international classification of disease, 10th revision (ICD-10) codes in emergency department data. Int J Popul Data Sci. 2018;3(1). doi:10.23889/ijpds.v3i1.445

10. Wong J, Abrahamowicz M, Buckeridge DL, Tamblyn R. Assessing the accuracy of using diagnostic codes from administrative data to infer antidepressant treatment indications: a validation study. Pharmacoepidemiol Drug Saf. 2018;27(10):1101–1111. doi:10.1002/pds.4436

11. Patel MR, Peterson ED, Dai D, Brennan M, Redberg R, Anderson V, et al. Low Diagnostic Yield of Elective Coronary Angiography. N Engl J Med. 2010;362(10):886–895. doi:10.1056/NEJMoa0907272

12. Harrison CJ, Sidey-Gibbons CJ. Machine learning in medicine: a practical introduction to natural language processing. BMC Med Res Methodol. 2021;21(1):158. doi:10.1186/s12874-021-01347-1

13. Olaronke I, Olaleke J. A Systematic Review of Natural Language Processing in Healthcare. Int J Inf Technol Comput Sci. 2015;08:44–50. doi:10.5815/ijitcs.2015.08.07

14. Clusmann J, Kolbinger FR, Muti HS, Carrero Z, Eckardt JN, Laleh NG et al. The future landscape of large language models in medicine. Commun Med. 2023;3(1):141. doi:10.1038/s43856-023-00370-1

15. Zhang X, Bellolio MF, Medrano-Gracia P, Werys K, Yang S, and Mahajan P. Use of natural language processing to improve predictive models for imaging utilization in children presenting to the emergency department. BMC Med Inform Decis Mak. 2019;19(1):287. doi:10.1186/s12911-019-1006-6

16. Myers D, Mohawesh R, Chellaboina VI, Sathvik AL, Venkatesh P, Ho YH, et al. Foundation and large language models: fundamentals, challenges, opportunities, and social impacts. Clust Comput. 2024;27(1):1–26. doi:10.1007/s10586-023-04203-7

17. Thirunavukarasu AJ, Ting DSJ, Elangovan K, Gutierrez L, Tan TF, and Shu Wei Ting D. Large language models in medicine. Nat Med. 2023;29(8):1930–1940. doi:10.1038/s41591-023-02448-8

18. Government of Canada SC. Population and Dwelling Count Highlight Tables, 2016 Census. Published February 8, 2017. Accessed March 25, 2024. https://www12.statcan.gc.ca/census-recensement/2016/dp-pd/hlt-fst/pd-pl/Comprehensive.cfm

19. Verma AA, Masoom H, Pou-Prom C, Shin S, Guerzhoy M, Fralick M et al. Developing and validating natural language processing algorithms for radiology reports compared to ICD-10 codes for identifying venous thromboembolism in hospitalized medical patients. Thromb Res. 2022;209:51–58. doi:10.1016/j.thromres.2021.11.020

20. Cohen J, Korevaar D, Altman D, Bruns D, Gatsonis C, Hooft L, et al. STARD 2015 guidelines for reporting diagnostic accuracy studies: explanation and elaboration. BMJ Open. 2016;6(11):e012799. doi:10.1136/bmjopen-2016-012799

21. Hersh WR, Weiner MG, Embi PJ, Logan JR, Payne P, Bernstam E, et al. Caveats for the Use of Operational Electronic Health Record Data in Comparative Effectiveness Research. Med Care. 2013;51:S30. doi:10.1097/MLR.0b013e31829b1dbd

22. Kore A, Abbasi Bavil E, Subasri V, Abdalla M, Fine B, Dolatabadi E, et al. Empirical data drift detection experiments on real-world medical imaging data. Nat Commun. 2024;15(1):1887. doi:10.1038/s41467-024-46142-w

23. Sandmann S, Riepenhausen S, Plagwitz L, and Varghese J. Systematic analysis of ChatGPT, Google search and Llama 2 for clinical decision support tasks. Nat Commun. 2024;15(1):2050. doi:10.1038/s41467-024-46411-8

24. Hu Y, Chen Q, Du J, Peng X, Keloth V, Zuo X, et al. Improving large language models for clinical named entity recognition via prompt engineering. J Am Med Inform Assoc. Published online January 27, 2024:ocad259. doi:10.1093/jamia/ocad259

25. Boussina, A., Krishnamoorthy, R., Quintero, K., Joshi S, Wardi G, Pour H, et al. (2024). Large Language Models for More Efficient Reporting of Hospital Quality Measures. NEJM AI, AIcs2400420.

26. Germini F, Hu Y, Afzal S, Al-haimus F, Puttagunta S, Niaz S. Feasibility of a quality improvement project to increase adherence to evidence-based pulmonary embolism diagnosis in the emergency department. Pilot Feasibility Stud. 2021;7(1):4. doi:10.1186/s40814-020-00741-8

27. Castiglioni I, Rundo L, Codari M, Di Leo G, Salvatore C, Interlenghi M, et al. AI applications to medical images: From machine learning to deep learning. Phys Medica Eur J Med Phys. 2021;83:9–24. doi:10.1016/j.ejmp.2021.02.006

28. Willemink MJ, Koszek WA, Hardell C, Wu J, Fleischmann D, Harvey H, et al. Preparing Medical Imaging Data for Machine Learning. Radiology. 2020;295(1):4–15. doi:10.1148/radiol.2020192224

29. Mullainathan S, Obermeyer Z. Diagnosing Physician Error: A Machine Learning Approach to Low-Value Health Care*. Q J Econ. 2022;137(2):679–727. doi:10.1093/qje/qjab046

30. Abonamah AA, Tariq MU, Shilbayeh S. On the Commoditization of Artificial Intelligence. Front Psychol. 2021;12. doi:10.3389/fpsyg.2021.696346

31. Wang J, Shi E, Yu S, Wu Z, Ma C, Dai M, et al. Prompt Engineering for Healthcare: Methodologies and Applications. arXiv.org. Published April 28, 2023. Accessed March 23, 2024. https://arxiv.org/abs/2304.14670v1

32. Li Y, Guo Y, Guerin F, and Lin C. Evaluating Large Language Models for Generalization and Robustness via Data Compression. Published online February 3, 2024. doi:10.48550/arXiv.2402.0086

